# Determinants of unmet social needs and the role of parental mental health in families from multicultural and regional/rural communities of Australia

**DOI:** 10.1101/2025.09.03.25335048

**Authors:** James John, Teresa Winata, Si Wang, Melissa Smead, Weng Tong Wu, Jane Kohlhoff, Virginia Schmied, Bin Jalaludin, Kenny Lawson, Siaw-Teng Liaw, Raghu Lingam, Andrew Page, Christa Lam-Cassettari, Katherine Boydell, Ping-I Lin, Ilan Katz, Ann Dadich, Shanti Raman, Rebekah Grace, Aunty Kerrie Doyle, Tom McClean, Blaise Di Mento, John Preddy, Susan Woolfenden, Valsamma Eapen

## Abstract

**Introduction:** Families from disadvantaged communities often experience social care needs that adversely impact access to social and healthcare services. This study aimed to explore the determinants of social care needs and the associated clinical characteristics such as parental mental health among families from multicultural and regional/rural communities of Australia.

**Methods:** This study is a secondary analysis of a randomised controlled trial conducted among parents/carers of children from culturally and linguistically diverse (CALD) communities of South Western Sydney and rural/regional communities of Murrumbidgee. The primary outcome of unmet social care needs was measured using the WE CARE survey. Binary logistic regression models were used to investigate the association between sociodemographic and clinical indicators associated with the risk of unmet needs at baseline (model 1), 6 months (model 2), and 12 months (model 3).

**Results:** Of the sample of 288 participants, 61% (n=176) reported one or more unmet needs. Findings of the regression analyses showed that clinical indicator such as parental mental distress (AOR 1.09, 95% CI 1.04, 1.16) alongside other sociodemographic factors such as CALD status (AOR 2.87, 95% CI 1.23, 6.69), lower levels parental education (AOR 5.76, 95% CI 2.21, 14.99), and marital status (AOR 2.07, 95% CI 1.01, 4.22) were associated with higher risk of unmet needs at baseline. Consistent with the baseline model, lower levels of parental education were significantly associated with two-to-three-fold higher odds of unmet needs at 6 and 12 months.

**Conclusion:** The study highlights the significant burden of unmet social needs among families from multicultural and rural/regional communities, emphasising the role of parental mental health and education levels as key contributing factors amongst other sociodemographic factors. Findings suggest the need for integrated, family-centred interventions that address both social and healthcare needs, particularly for vulnerable populations.

## Introduction

Unmet social care needs refer to the absence of critical resources such as housing, food security, employment, and access to mental and physical healthcare [1]. These challenges are exacerbated among families from priority population groups such as those from culturally and linguistically diverse (CALD) as well as regional or rural communities, where systemic barriers and socioeconomic disparities often limit access to social and healthcare services. In Australia, multicultural families, including migrants and refugees, face unique challenges due to language barriers, cultural adaptation, and discrimination, which can hinder their ability to access services [2, 3]. Similarly, families in regional and rural areas encounter inequity in access due to geographic location, fewer healthcare facilities, and limited employment opportunities, exacerbating their social and economic vulnerabilities [4, 5]. Hence, addressing these unmet needs is crucial, as they are strongly linked to poorer physical and mental health outcomes, intergenerational disadvantage, and reduced quality of life.

Australian health and social policies have emphasised the need for integrated, community-based approaches that address both medical and social determinants of health [6–9]. Despite these efforts, gaps persist in reaching high-risk populations, particularly those from the priority population groups. Existing service models often operate in silos, failing to provide comprehensive, coordinated care that addresses the multifaceted needs of families facing social disadvantage [10, 11]. The social determinants of health (SDOH) framework provides a comprehensive approach for understanding how social, economic, and environmental factors collectively shape health outcomes. Studies have demonstrated the effectiveness of early intervention programs that address social determinants of health, leading to improved health and wellbeing and cost beneficial outcomes [12, 13]. However, there is limited research on the specific factors and extent of unmet social needs and the interaction with parental mental health status and child developmental concerns among multicultural and regional/rural families in Australia.

To address this knowledge gap, this study aimed to determine the sociodemographic, sociocultural, and clinical indicators such as child developmental concerns and parental mental health associated with unmet social needs among families from multicultural and regional/rural communities of Australia. Findings of this study will provide critical insights into the barriers these families face in accessing essential social and healthcare supports. The evidence generated will inform targeted policy reforms and the development of interventions aimed at enhancing service integration and accessibility. Further, identifying and addressing these critical social care needs is crucial for promoting social equity, improving health outcomes, and ensuring that all Australian families have the support they need to thrive, regardless of their cultural, socioeconomic, and geographic background.

## Methods

### Study design and participants

Study findings are reported following the Consolidated Standards of Reporting Trials (CONSORT) guidelines [14]. This study is a secondary analysis of data from a parallel group randomised controlled trial (RCT) conducted among two priority population communities of South Western Sydney Local Health District (SWSLHD) and Murrumbidgee Local Health District (MLHD) representing predominantly multicultural/low-income and rural/regional communities, respectively. The authors confirm that all ongoing and related trials for this intervention are registered (ACTRN12621000766819). Parents/caregivers with a child aged six months to three years old attending Child and Family Health Services (CFHS) and other services providing child and family health care were recruited to participate in this study. All participants provided written informed consent. The recruitment and follow-up period of this study was between 1 August 2021 and 30 June 2023 with the recruitment commencing at the same time across both study sites. Further information on the randomisation and blinding, sample size calculation, and the intervention components are detailed in the published protocol [15].

### Outcome measure - Unmet social needs

Unmet social care needs in this study were assessed using the WE CARE survey, a brief and validated screening tool designed to efficiently identify the presence and extent of unmet social needs in families [16]. The survey comprises six binary (yes/no) responses related to family psychosocial needs, covering areas such as childcare, employment, homelessness, food security, education, and utilities. Additional questions were included for participants who respond affirmatively to any of the initial six items, prompting participants to indicate, on a three-point scale (yes, no, or maybe later), whether they require further assistance in addressing their unmet psychosocial needs. For the purpose of this study, the presence of unmet needs was dichotomised as no unmet needs (WE CARE score = 0) and one or more unmet needs (WE CARE score ≥ 1).

### Explanatory variables

The clinical and health service use indicators included number of concerns reported via the Learn the Signs Act Early (LTSAE) assessment, parental mental health assessed via Kessler’s psychological distress scale (K10), and current service use (no, yes). The LTSAE is developed by the Centres for Disease Control and Prevention (CDC) [17], to monitor child’s development from 2 months to 5 years old. The assessment includes several domains of social and emotional, language/communication, cognitive, and movement/physical development. The K10 assessment is a widely used self-report questionnaire designed to measure psychological distress in individuals [18]. It comprises 10 items with ratings on a five-point scale assessing various aspects of emotional wellbeing and mental health over the past four weeks.

The sociodemographic and sociocultural factors included: child’s age (in months), child’s gender (female, male), parent’s age (in years), CALD status (yes, no), highest level of education (Postgraduate/graduate diploma/ Bachelors, Advance diploma/Certificate ¾, Year 12 and/or below), marital status (Married, others – de facto/single/divorced), IRSAD quintiles (from Q1 - most disadvantaged to Q5 - the most advantaged), and treatment group (intervention, control).

### Statistical analysis

Descriptive characteristics of the sample was computed using mean with standard deviation (SD) for continuous variables and frequency counts with percentages for categorical variables. Shapiro-Wilks test for normality and analysis of normal quantile-quantile plots were used to assess the normality of distribution. Univariate linear regression analysis was used to determine the factors independently associated with the risk of unmet needs. Pearson correlation was conducted to explore any significant correlation between the variables of interest. Primary analysis included three binary logistic regression analysis models to determine the sociodemographic, sociocultural, and clinical indicators associated with unmet social needs at baseline (model 1), 6 months (model 2), and 12 months (model 3). All analyses were undertaken in Statistical Package for the Social Sciences (SPSS) version 28 (IBM SPSS, IBM Corp., Armonk, NY, USA)

## Results

### Participant characteristics

The CONSORT flowchart showing randomisation of participants along with follow-up at different time points is summarised in **Figure 1**. The descriptive characteristics of the sample at baseline by unmet needs status is presented in **Table 1**. Of the total sample of 288 participants, 61% (n=176) reported one or more unmet needs at baseline with the top concerns being help with food, mental health, employment, and day care.

**Figure 1.**
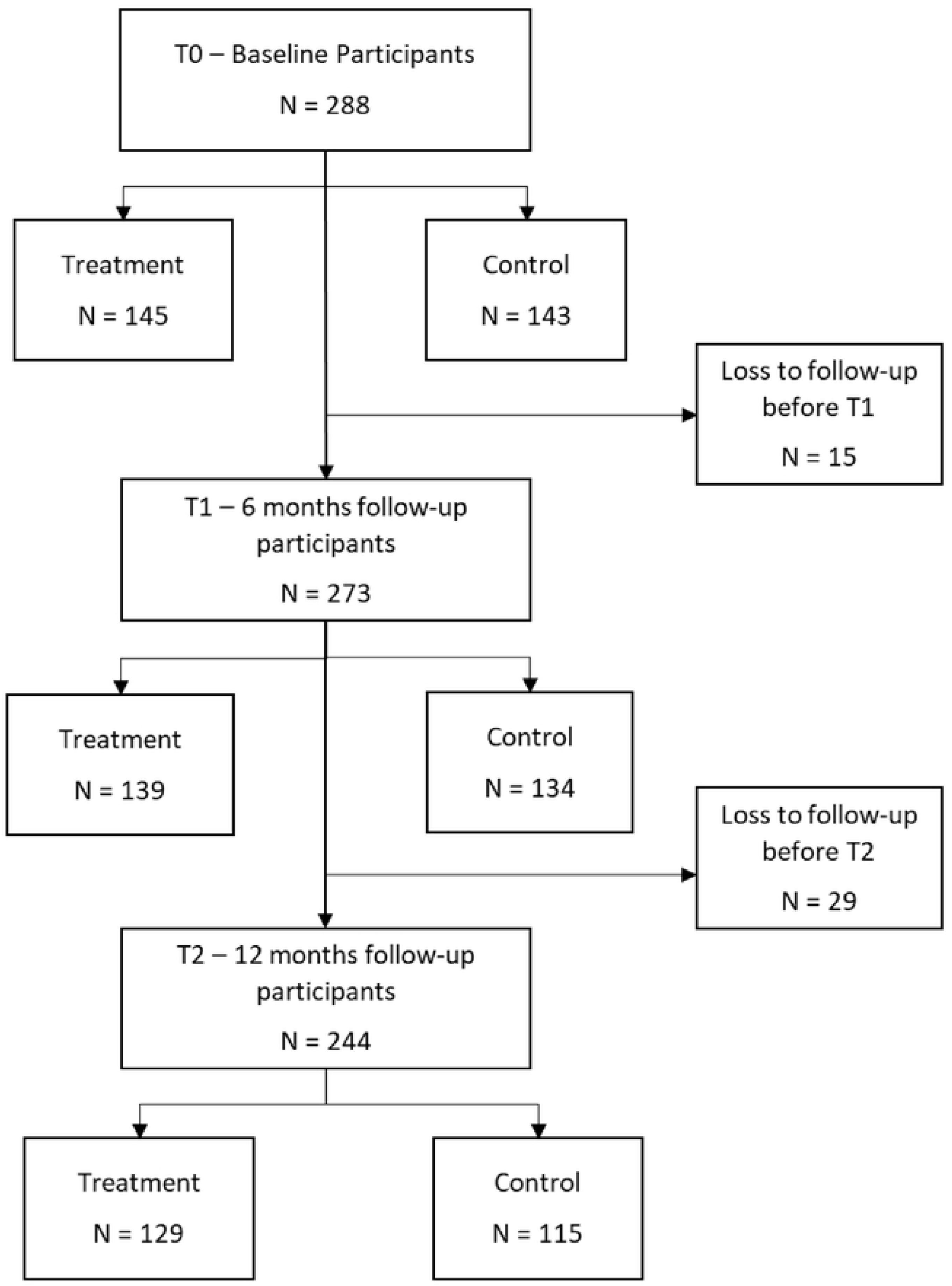
CONSORT study Flow Diagram.

**Table 1.** Descriptive characteristics of participants at baseline.

| **Variables** | **Total sample**<br>**(N=288)**<br>**n (%) or M(SD)** | **No unmet needs**<br>**(N=112)**<br>**n (%) or M(SD)** | **One or more unmet needs**<br>**(N=176)**<br>**n (%) or M(SD)** |
| --- | --- | --- | --- |
| **Sociodemographic and sociocultural factors** |  |  |  |
| Child's age in months, mean (SD) | 18.84 (9.42) | 18.61 (9.10) | 18.98 (9.64) |
| Child's gender |  |  |  |
| *Female* | 145 (50.3) | 58 (51.8) | 87 (49.4) |
| *Male* | 143 (49.7) | 54 (48.2) | 89 (50.6) |
| Parent/Caregiver's age in years, mean (SD) | 31.23 (6.50) | 31.96 (5.53) | 30.77 (7.03) |
| CALD status<sup>#</sup> |  |  |  |
| *No* | 206 (71.5) | 87 (77.7) | 119 (67.6) |
| *Yes* | 46 (16.0) | 16 (14.3) | 30 (17.0) |
| *Missing* | 62 (21.5) | 9 (8.0) | 27 (15.3) |
| Parent/Caregiver's education level |  |  |  |
| *Postgraduate/Graduate diploma/Bachelor's degree* | 148 (51.4) | 71 (63.4) | 77 (43.8) |
| *Advanced Diploma/ Certificate III/IV* | 78 (27.1) | 33 (29.5) | 45 (25.6) |
| *Up to Year 12* | 62 (21.5) | 8 (7.1) | 54 (30.7) |
| **Marital status** |  |  |  |
| *Married* | 178 (61.8) | 85 (75.9) | 93 (52.8) |
| *De facto/Single/Divorced* | 109 (37.8) | 27 (24.1) | 82 (46.6) |
| *Missing* | 1 (0.3) | 0 (0.0) | 1 (0.6) |
| **Socioeconomic status (IRSAD)** |  |  |  |
| *Quintiles 1-2 (disadvantaged)* | 160 (55.6) | 65 (58.0) | 95 (54.0) |
| *Quintile 3* | 109 (37.8) | 37 (33.0) | 72 (40.9) |
| *Quintiles 4-5 (advantaged)* | 19 (6.6) | 10 (8.9) | 9 (5.1) |
| **Treatment group** |  |  |  |
| *Control* | 143 (49.7) | 45 (40.2) | 98 (55.7) |
| *Intervention* | 145 (50.3) | 67 (59.8) | 78 (44.3) |
| **Clinical indicators and health service use** |  |  |  |
| No of child developmental concerns, mean (SD) | 0.27 (0.66) | 0.13 (0.44) | 0.36 (0.76) |
| Parental mental wellbeing (K10 scores), mean (SD) | 18.47 (6.78) | 16.19 (4.32) | 19.92 (7.62) |
| Premature birth |  |  |  |
| *No* | 272 (94.4) | 107 (95.5) | 165 (93.8) |
| *Yes* | 16 (5.6) | 5 (4.5) | 11 (6.3) |
| Current service use |  |  |  |
| *No* | 211 (73.3) | 80 (71.4) | 131 (74.4) |
| *Yes* | 73 (25.3) | 30 (26.8) | 43 (24.4) |
| *Missing* | 4 (1.4) | 2 (1.8) | 2 (1.1) |

#CALD status – parent’s country of birth and/or preferred language

### Findings of the regression analyses

Findings of the univariate regression analysis (**Table 2**) showed significant associations between clinical indicators such as parental mental health issues and number of child developmental concerns, as well as sociodemographic factors such as lower level of parental education (up to year 12) and marital status (de facto, single, or divorced) to be significantly associated with higher odds of unmet needs whereas those in the intervention group had lower odds of unmet social needs.

**Table 2.** Associations between sociodemographic factors, clinical factors, and unmet social needs (unadjusted models)

| Variables | At baseline | At 6 months | At 12 months |
| --- | --- | --- | --- |
|  | COR (95% CI) | COR (95% CI) | COR (95% CI) |
| Child's age in months, mean (SD) | 1.00 (0.97, 1.03) | 1.01 (0.99, 1.04) | 0.99 (0.97, 1.02) |
| Child's gender |  |  |  |
| *Female* | Reference category | Reference category | Reference category |
| *Male* | 1.09 (0.68, 1.77) | 1.09 (0.67, 1.75) | 0.85 (0.51, 1.40) |
| Parent/Caregiver's age in years, mean (SD) | 0.97 (0.94, 1.01) | 0.99 (0.96, 1.03) | 0.96 (0.93, 1.01) |
| CALD status<sup>#</sup> |  |  |  |
| *No* | Reference category | Reference category | Reference category |
| *Yes* | 1.37 (0.70, 2.67) | 1.12 (0.58, 2.17) | 0.92 (0.45, 1.86) |
| Parent/Caregiver's education level |  |  |  |
| *Postgraduate/Graduate diploma/Bachelor's degree* | Reference category | Reference category | Reference category |
| *Advanced Diploma/ Certificate III/IV* | 1.26 (0.72, 2.19) | 1.36 (0.77, 2.41) | 1.48 (0.81, 2.70) |
| *Up to Year 12* | 6.22 (2.77, 13.98)* | 2.70 (1.39, 5.24)* | 3.04 (1.53, 6.04)* |
| Marital status |  |  |  |
| *Married* | Reference category | Reference category | Reference category |
| *De facto/Single/Divorced* | 2.78 (1.64, 4.69)* | 1.26 (0.77, 2.07) | 2.04 (1.20, 3.47)* |
| Socioeconomic status (IRSAD) |  |  |  |
| *Quintiles 1-2 (disadvantaged)* | Reference category | Reference category | Reference category |
| *Quintile 3* | 1.33 (0.80, 2.21) | 1.11 (0.67, 1.83) | 1.36 (0.81, 2.30) |
| *Quintiles 4-5 (advantaged)* | 0.61 (0.24, 1.59) | 1.17 (0.42, 3.23) | 0.69 (0.22, 2.23) |
| Treatment group |  |  |  |
| *Control* | Reference category | Reference category | Reference category |
| *Intervention* | 0.54 (0.33, 0.87)* | 0.63 (0.39, 1.03) | 0.61 (0.37, 0.99)* |
| **Clinical indicators and health service use at baseline** |  |  |  |
| No of child developmental concerns, mean (SD) | 1.97 (1.21, 3.21)* | 1.72 (1.10, 2.69)* | 1.92 (1.19, 3.08)* |
| Parental mental wellbeing (K10 scores), mean (SD) | 1.10 (1.05, 1.15)* | 1.03 (0.99, 1.07) | 1.06 (1.02, 1.11)* |
| Premature birth |  |  |  |
| *No* | Reference category | Reference category | Reference category |
| *Yes* | 1.43 (0.48, 4.22) | 1.32 (0.47, 3.73) | 1.64 (0.52, 5.17) |
| Current service use |  |  |  |

| *No* | Reference category | Reference category | Reference category |
| --- | --- | --- | --- |
| *Yes* | 0.88 (0.51, 1.51) | 1.02 (0.59, 1.78) | 1.24 (0.70, 2.19) |

COR – Crude Odds Ratio; CI – Confidence Intervals; \*p-value<0.05

Consistent with the univariate regression analysis, the multivariate binary logistic regression models (**Table 3**) showed that higher scores on parental mental health issues via K10 were associated with higher odds of unmet needs at baseline (AOR 1.09, 95% CI: 1.04, 1.16). Child developmental concerns did not retain significance in the adjusted models. Further, current service use showed no significant main effect, nor did it significantly moderate the association between K10, child developmental concerns, and unmet needs.

**Table 3.** Associations between sociodemographic factors, clinical factors, and unmet social needs (adjusted models)

|  | At baseline | At 6 months | At 12 months |
| --- | --- | --- | --- |
| Variables | AOR (95% CI) | AOR (95% CI) | AOR (95% CI) |
| Child's age in months, mean (SD) | 0.99 (0.96, 1.02) | 1.01 (0.98, 1.04) | 0.98 (0.95, 1.02) |
| Child's gender |  |  |  |
| *Female* | Reference category | Reference category | Reference category |
| *Male* | 1.01 (0.56, 1.83) | 1.12 (0.65, 1.95) | 0.70 (0.39, 1.27) |
| Parent/Caregiver's age in years, mean (SD) | 1.01 (0.95, 1.08) | 1.02 (0.96, 1.07) | 1.02 (0.96, 1.08) |
| CALD status<sup>#</sup> |  |  |  |
| *No* | Reference category | Reference category | Reference category |
| *Yes* | 2.87 (1.23, 6.69)* | 1.22 (0.57, 2.63) | 1.53 (0.67, 3.52) |
| Parent/Caregiver's education level |  |  |  |
| *Postgraduate/Graduate diploma/Bachelor's degree* | Reference category | Reference category | Reference category |
| *Advanced Diploma/ Certificate III/IV* | 1.31 (0.64, 2.66) | 1.37 (0.69, 2.69) | 1.29 (0.62, 2.69) |
| *Up to Year 12* | 5.76 (2.21, 14.99)* | 3.43 (1.52, 7.72)* | 2.52 (1.05, 6.06)* |
| Marital status |  |  |  |
| *Married* | Reference category | Reference category | Reference category |
| *De facto/Single/Divorced* | 2.07 (1.01, 4.22)* | 1.25 (0.65, 2.37) | 1.84 (0.91, 3.73) |
| Socioeconomic status (IRSAD) |  |  |  |
| *Quintiles 1-2 (disadvantaged)* | Reference category | Reference category | Reference category |
| *Quintile 3* | 1.33 (0.68, 2.59) | 1.12 (0.61, 2.07) | 1.21 (0.62, 2.35) |
| *Quintiles 4-5 (advantaged)* | 0.50 (0.15, 1.72) | 2.11 (0.68, 6.58) | 0.76 (0.19, 3.73) |
| Treatment group |  |  |  |
| *Control* | Reference category | Reference category | Reference category |
| *Intervention* | 0.32 (0.17, 0.59)* | 0.59 (0.34, 1.04) | 0.49 (0.27, 0.92)* |
| **Clinical indicators and health service use at baseline** |  |  |  |
| No of child developmental concerns, mean (SD) | 1.68 (0.87, 3.24) | 1.37 (0.81, 2.31) | 1.45 (0.84, 2.53) |
| Parental mental wellbeing (K10 scores), mean (SD) | 1.09 (1.04, 1.16)* | 1.01 (0.96, 1.05) | 1.04 (0.99, 1.09) |
| Premature birth |  |  |  |
| *No* | Reference category | Reference category | Reference category |
| *Yes* | 1.21 (0.30, 4.86) | 1.14 (0.33, 4.01) | 3.16 (0.75, 13.29) |
| Current service use |  |  |  |

| *No* | Reference category | Reference category | Reference category |
| --- | --- | --- | --- |
| *Yes* | 0.74 (0.37, 1.46) | 1.10 (0.59, 2.05) | 1.23 (0.63, 2.39) |

AOR – Adjusted Odds Ratio; CI – Confidence Intervals; \*p-value<0.05

In terms of sociodemographic and sociocultural factors, those from a CALD background had more than double the odds of unmet needs at baseline (AOR 2.87, 95% CI: 1.23, 6.69). Lower level of parental education (up to year 12) were associated with six-time higher odds of having one or more unmet needs (AOR 5.76, 95% CI: 2.21, 14.99) at baseline compared to those with tertiary level education. Compared to parents/caregivers who were married, those who were de facto, single, or divorced had double the odds of having one or more unmet needs (AOR 2.07, 95% CI: 1.01, 4.22) at baseline. Similar to model 1 (baseline), lower levels of parental education were significantly associated with two-to-three-fold higher odds of unmet needs at 6 and 12 months (models 2 and 3). On the other hand, compared to those in the control group, participants in the intervention group had lower odds of unmet needs both at baseline (AOR 0.32, 95% CI: 0.17, 0.59) and at 12 months (AOR 0.49, 95% CI: 0.27, 0.92). Current service use did not moderate the association between CALD, parental education, marital status, and unmet needs.

## Discussion

This study aimed to identify the sociodemographic, sociocultural, and clinical indicators of unmet social needs among families from CALD and regional/rural communities in Australia. Our findings highlighted that a substantial proportion (61%) of families reported at least one unmet social need at baseline, with food insecurity, employment, and daycare needs being the most prevalent concerns. Additionally, parental mental health issues and key social and clinical risk factors such as CALD status, parental education, and marital status were found to be significantly associated with unmet needs. This underscores the urgent need for targeted, culturally sensitive, and multifaceted interventions and policies to effectively address these complex social challenges and improve the wellbeing of vulnerable families across Australia.

Poor parental mental health was a critical determinant and was associated with increased risk of unmet needs. There is substantial evidence highlighting the bidirectional relationship where poor mental health can both contribute to and result from social disadvantage [19, 20]. Parents experiencing psychological distress, including anxiety and depression, often face difficulties in fulfilling their caregiving roles effectively. Poor mental health can compromise their ability to meet their children’s basic needs, maintain stable housing, or navigate social and healthcare systems, thereby compounding existing social vulnerabilities [20–22]. Consequently, examining the role of parental mental health in predicting social disparities is vital for designing effective interventions. Understanding how poor mental health drives unmet social needs can inform the development of holistic, wraparound interventions that addresses both psychological wellbeing and social determinants concurrently, thereby reducing disparities and improving family outcomes over the long term.

Consistent with previous research, lower parental education emerged as one of the strongest predictors of unmet social needs, with those having only up to year 12 education facing six- fold greater odds of unmet needs compared to parents with tertiary education. This highlights the critical role of education in providing families with health literacy, problem-solving, and access to services that can mitigate social vulnerabilities [23]. Further, it is well established that higher education is strongly linked to better employment opportunities and income stability, further reducing the risk of social disadvantage [24]. Consequently, policies and interventions aimed at reducing unmet social needs must prioritise addressing educational inequalities such as providing access and support to health literacy and community learning programs. By indirectly strengthening families’ resources and capacities through education, these initiatives have the potential to significantly alleviate social vulnerabilities and improve overall family outcomes [25].

We also found key sociocultural factors such as family structure and CALD status to further influence the extent to which families experience unmet social needs. Marital status was a significant factor, where parents who were single, divorced, or in de facto relationships had about twice the odds of experiencing unmet needs compared to married parents. This finding aligns with literature suggesting that single-parent and non-traditional family structures often face greater financial and social pressures, which may limit their ability to meet essential needs [26–28]. Notably, families from CALD backgrounds experienced nearly three times higher odds of unmet needs, reiterating the substantial barriers faced by those from CALD communities. These barriers may include language difficulties, lack of familiarity with available services, and potential discrimination, restricting access to essential supports [2, 29, 30]. These findings highlight the need for culturally sensitive, family-centred policies and support systems that address the specific challenges faced by diverse family structures and CALD communities to reduce social inequities and improve access to critical resources.

### Strengths, Limitations, Directions for future research

This study has several strengths and limitations. One of the key strengths of this research is that it addresses a gap in understanding the determinants of unmet social needs in multicultural and regional/rural Australian families, populations often underrepresented in research. The use of validated measures for mental health and unmet needs enhances the reliability of findings.

However, several limitations should be acknowledged. The reliance on self-reported data may introduce reporting bias or social desirability effects. Our sample, although diverse, may not fully represent all multicultural or regional/rural groups across Australia, limiting generalisability.

Future studies should aim to explore specific cultural, linguistic, and geographic barriers via qualitative research in greater detail to inform more nuanced and effective interventions for multicultural and regional/rural communities. Evaluations of digital and community-based navigation models over longer periods will help clarify optimal implementation strategies and sustainability. Finally, investigating policy-level interventions addressing structural determinants, such as housing and employment policies, will be crucial to comprehensively reducing unmet social needs.

### Implications for Policy and Practice

Our findings highlight the importance of tailored approaches to identify and address unmet social needs among vulnerable families. Policies should prioritise education as a key social determinant, integrating education and family support programs to empower parents. Mental health services should be embedded within community and social care frameworks, with culturally sensitive approaches and virtual support to meet the specific needs of CALD and rural/regional populations, respectively. Family support services must recognise individual family dynamics such as the additional challenges faced by single-parent and non-traditional families, ensuring access to flexible and affordable childcare, employment support, and food assistance. The demonstrated potential of digital screening and navigation interventions suggests that scaling such models could improve service access and reduce unmet needs, particularly when adapted for cultural and regional contexts.

## Conclusion

Our study provides valuable insights into the key risk factors associated with the risk of unmet social needs among families from diverse CALD and regional/rural communities in Australia that often leads to inequity in access to services. The findings emphasise the complex and interconnected nature of sociodemographic, sociocultural, clinical factors that contribute to social vulnerabilities within these priority populations. It is imperative that policies and programs adopt a holistic, culturally sensitive approach that addresses social disparities and integrates mental health supports to overcome barriers to service access, in particular for culturally and linguistically diverse families. Further research and sustained investment in tailored interventions are critical to promoting equity and wellbeing for all Australian families.

## Data Availability

Data are available upon reasonable request. Data collected for this study will be shared upon reasonable request to the corresponding author

## Declarations

### Data sharing statement

Data are available upon reasonable request. Data collected for this study will be shared upon reasonable request.

### Human Ethics and Consent to Participate Declarations

The study conforms to the principles outlined in the Declaration of Helsinki. All methods were carried out in accordance with relevant guidelines and regulations of The National Statement on Ethical Conduct in Human Research (2023). The South Western Sydney Local Health District Human Research Ethics Committee approved this study (2020/ETH01418). All participating parents have provided written informed consent prior to participation.

### Author Contribution

Conceptualisation: VE, JP, and SW; Data curation: JJ, TW, SW, and MS; Methodology and Formal analysis: JJ and TW; Funding acquisition: VE, JP, SW, JK, VS, BJ, KL, S-TL, RL, AP, KB, P-IL, IK, AD, SR, RG, AKD, and TM; Project administration: TW, SW, and MS; Writing. – original draft: JJ and TW; Writing – review & editing: All authors.

## References

1. Garg A, Marino M, Vikani AR, Solomon BS. Addressing families’ unmet social needs within pediatric primary care: the health leads model. Clinical pediatrics. 2012;51(12):1191–3.

2. Khatri RB, Assefa Y. Access to health services among culturally and linguistically diverse populations in the Australian universal health care system: issues and challenges. BMC Public Health. 2022;22(1):880. doi: 10.1186/s12889-022-13256-z.

3. Ng ZY, Waite M, Hickson L, Ekberg K. Language accessibility in allied healthcare for culturally and linguistically diverse (CALD) families of young children with chronic health conditions: a qualitative systematic review. Speech, Language and Hearing. 2021;24(2):50–66.

4. Kavanagh BE, Corney KB, Beks H, Williams LJ, Quirk SE, Versace VL. A scoping review of the barriers and facilitators to accessing and utilising mental health services across regional, rural, and remote Australia. BMC health services research. 2023;23(1):1060.

5. Thorn H, Olley R. Barriers and facilitators to accessing medical services in rural and remote Australia: a systematic review. Asia Pacific Journal of Health Management. 2023;18(1):20–9.

6. National Mental Health Commission. National Children’s Mental Health and Wellbeing Strategy 2021. 2021.

7. Productivity Commission. PC Productivity Insights: Australia’s long term productivity experience. 2020.

8. NSW Health. The First 2000 Days Framework. 2019.

9. Health. AGDo. National action plan for the health of children and young people: 2020– 2030. 2019.

10. Psaila K, Schmied V, Fowler C, Kruske S. Discontinuities between maternity and child and family health services: health professional’s perceptions. BMC health services research. 2014;14:1–12.

11. Schmied V, Homer C, Fowler C, Psaila K, Barclay L, Wilson I, et al. Implementing a national approach to universal child and family health services in A ustralia: professionals’ views of the challenges and opportunities. Health & Social Care in the Community. 2015;23(2):159–70.

12. Whitman A, De Lew N, Chappel A, Aysola V, Zuckerman R, Sommers BD. Addressing social determinants of health: Examples of successful evidence-based strategies and current federal efforts. Off Heal Policy. 2022;1:1–30.

13. Holt-Lunstad J. Social connection as a public health issue: The evidence and a systemic framework for prioritizing the “social” in social determinants of health. Annual Review of Public Health. 2022;43(1):193–213.

14. Hopewell S, Chan A-W, Collins GS, Hróbjartsson A, Moher D, Schulz KF, et al. CONSORT 2025 statement: updated guideline for reporting randomised trials. The Lancet. 2025;405(10489):1633–40.

15. Eapen V, Woolfenden S, Schmied V, Jalaludin B, Lawson K, Liaw S, et al. “Watch Me Grow-Electronic (WMG-E)” surveillance approach to identify and address child development, parental mental health, and psychosocial needs: study protocol. BMC Health Services Research. 2021;21:1–10.

16. Garg A, Butz AM, Dworkin PH, Lewis RA, Thompson RE, Serwint JR. Improving the management of family psychosocial problems at low-income children’s well-child care visits: the WE CARE Project. Pediatrics. 2007;120(3):547–58.

17. Moore T, Arefadib N, Deery A, Keyes M, West S. The first thousand days: an evidence paper. Melbourne: Murdoch Children’s Research Institute. 2017.

18. Kessler RC, Barker PR, Colpe LJ, Epstein JF, Gfroerer JC, Hiripi E, et al. Screening for Serious Mental Illness in the General Population. Archives of General Psychiatry. 2003;60(2):184–9. doi: 10.1001/archpsyc.60.2.184 %J Archives of General Psychiatry.

19. Mezzina R, Gopikumar V, Jenkins J, Saraceno B, Sashidharan S. Social vulnerability and mental health inequalities in the “Syndemic”: Call for action. Frontiers in psychiatry. 2022;13:894370.

20. McLoyd VC, Wilson L. The strain of living poor: Parenting, social support, and child mental health. Children in poverty: Child development and public policy. 1991:105–35.

21. Johnson SE, Lawrence D, Perales F, Baxter J, Zubrick SR. Poverty, parental mental health and child/adolescent mental disorders: Findings from a national Australian survey. Child Indicators Research. 2019;12(3):963–88.

22. Taylor M, Stevens G, Agho K, Raphael B. The impacts of household financial stress, resilience, social support, and other adversities on the psychological distress of Western Sydney parents. International Journal of Population Research. 2017;2017(1):6310683.

23. Mastekaasa A, Birkelund GE. The intergenerational transmission of social advantage and disadvantage: comprehensive evidence on the association of parents’ and children’s educational attainments, class, earnings, and status. European Societies. 2023;25(1):66–86.

24. Erola J, Jalonen S, Lehti H. Parental education, class and income over early life course and children’s achievement. Research in Social Stratification and Mobility. 2016;44:33–43.

25. Nutbeam D, McGill B, Premkumar P. Improving health literacy in community populations: a review of progress. Health promotion international. 2018;33(5):901–11.

26. Crosier T, Butterworth P, Rodgers B. Mental health problems among single and partnered mothers: The role of financial hardship and social support. Social psychiatry and psychiatric epidemiology. 2007;42(1):6–13.

27. Stack RJ, Meredith A. The impact of financial hardship on single parents: An exploration of the journey from social distress to seeking help. Journal of family and economic issues. 2018;39(2):233–42.

28. Martin MA. Family structure and the intergenerational transmission of educational advantage. Social science research. 2012;41(1):33–47.

29. Filia K, Menssink J, Gao CX, Rickwood D, Hamilton M, Hetrick S, et al. Social inclusion, intersectionality, and profiles of vulnerable groups of young people seeking mental health support. Social psychiatry and psychiatric epidemiology. 2022;57(2):245–54.

30. Smith C, Boylen S, Mutch R, Cherian S. Hear our voice: Pediatric communication barriers from the perspectives of refugee mothers with limited English proficiency. Journal of Pediatric Health Care. 2024;38(2):114–26.

